# Declining SARS-CoV-2 PCR sensitivity with time and dependence on clinical features: consequences for control

**DOI:** 10.1101/2020.08.23.20179408

**Authors:** B.J.M. Bergmans, C.B.E.M. Reusken, A.J.G. van Oudheusden, G.J. Godeke, A.A. Bonačić Marinović, E. de Vries, Y.C.M. Kluiters-de Hingh, R. Vingerhoets, M.A.H. Berrevoets, J.J. Verweij, A.E. Nieman, J. Reimerink, J.L. Murk, A.N. Swart

**Author notes:** Correspondence to: A.N. Swart.

## Abstract

Real-time reverse transcription-polymerase chain reaction (RT-PCR) on upper respiratory tract (URT) samples is the primary method to diagnose SARS-CoV-2 infections and guide public health measures, with a supportive role for serology. However, the clinical sensitivity of RT-PCR remains uncertain. In the present study, Bayesian statistical modeling was used to retrospectively determine the sensitivity of RT-PCR using SARS-CoV-2 serology in 644 COVID-19-suspected patients with varying degrees of disease severity and duration. The sensitivity of RTPCR ranged between 79–95%; while increasing with disease severity, it decreased rapidly over time in mild COVID-19 cases. Negative URT RT-PCR results should therefore be interpreted in the context of clinical characteristics, especially with regard to containment of viral transmission based on the ‘test, trace and isolate’ principle.

## Introduction

COVID-19 is diagnosed primarily by testing upper respiratory tract (URT) samples with real-time reverse transcription-polymerase chain reaction (RT-PCR)(*1*). Experience with nucleic acid amplification tests for other respiratory viruses, such as influenza virus, granted a high level of confidence in the clinical sensitivity of these types of assays(*2*). Early in the SARS-CoV-2 pandemic, however, Chinese clinicians already reported substantial numbers of negative tests of URT samples from patients with a high level of clinical suspicion for COVID-19(*3, 4*). Frequent false-negatives were later also reported by other physicians worldwide(*5, 6*) and were indicated to significantly complicate healthcare organization, hospital admission and isolation capacity (*7, 8*).

In several systematic reviews, the false-negative rate of RT-PCR was calculated to range between 22–66%, depending on symptom duration(*9, 10*). It is not known whether false-negatives were due to methodological problems, such as sampling error, suboptimal handling of samples or suboptimal assay design, or if they reflect a biological feature of SARS-CoV-2 infections. Clinical samples of lower respiratory tract material such as sputum or broncho-alveolar lavage fluid (BALF) appeared to yield significantly lower false negative rates than URT samples in patients with COVID-19 pneumonia (*3, 11, 12*). However, only 13–30% of patients produce sputum(*7*) and obtaining BALF is often impossible due to severe respiratory distress, or not indicated due to mild disease. URT samples are therefore the preferred and most feasible sample type. Estimates of sensitivity are further complicated by the lack of a reliable gold standard. Although imaging techniques may aid in the diagnosis of COVID-19(*13, 14*), their sensitivity and specificity are insufficient to be used as such.

Although complex, it is crucial to reliably assess the sensitivity of RT-PCR in different clinical cohorts as RT-PCR is the foundation for test, trace and isolation policies that are the cornerstone of worldwide pandemic control efforts. Highly sensitive and specific antibody assays are important tools to support patient diagnostics at a later stage of the disease. We postulate these can also be used to investigate the clinical sensitivity of RT-PCR. In the present study, Bayesian statistical modeling was used to retrospectively determine the clinical sensitivity of RT-PCR in URT-samples from a large cohort of COVID-19-suspected patients with varying degrees of disease severity and symptom duration using serology assays.

## Results

In the study period, 644 patients (>18 years of age) that presented at our hospital with clinically suspected COVID-19 were tested for SARS-CoV-2 by RT-PCR on URT samples. URT material was obtained by combined swabbing of subsequentially oropharynx (OP) and nasopharynx (NP), using the same swab, to increase sensitivity and reduce the logistic and resource burden(*15*). In addition, serum samples were taken after a minimum of 12 days post disease onset whenever possible, to determine if specific antibodies to SARS-CoV-2 had developed. Samples were obtained either through collection of left-over serum, taken for other purposes (N = 109) or by asking patients to have a blood sample taken for the purpose of this study (N = 141). In total, sera from 250 patients were obtained and analyzed for the presence of SARS-CoV-2 specific antibodies (Table 1, Fig. 1). To this goal, two independent tests were used; an in-house developed protein micro-array based on SARS-CoV-2 S1 and N proteins (PMA (*16, 17*)) and the Wantai total antibody ELISA based on the RBD-domain of S1 (Beijing Wantai Biological Pharmacy Enterprise, Beijing, China; Cat # WS1096). The manufacturers of the Wantai assay recommend index values of 1.1 and higher as evidence for the presence of specific antibodies. However, we observed that at this cut-off value, several RT-PCR confirmed patients scored negative and a cut-off of 0.25 was calculated to be more appropriate in our cohort (Fig. 2, Supplementary Text). The rightmost panel of this figure gives quantitative evidence. The histogram is scaled to 100% height per bar, and colored according to percentage of PCR positive (purple) and negative (yellow) status of the patients. Clearly, using the manufacturer’s cut-off, many positives would be to the left of the cut-off and would score false-negative in the Wantai test.

**Table 1.**
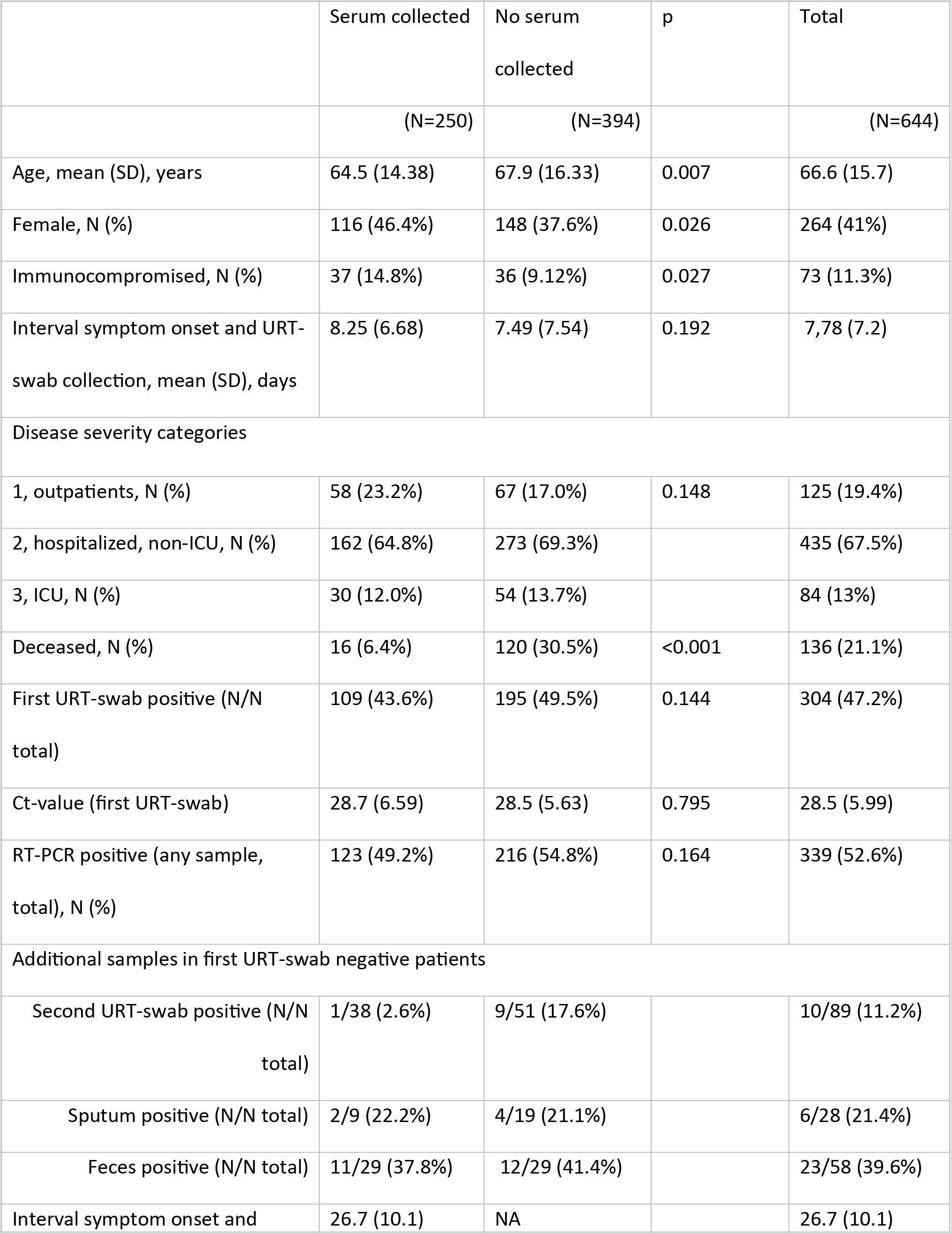
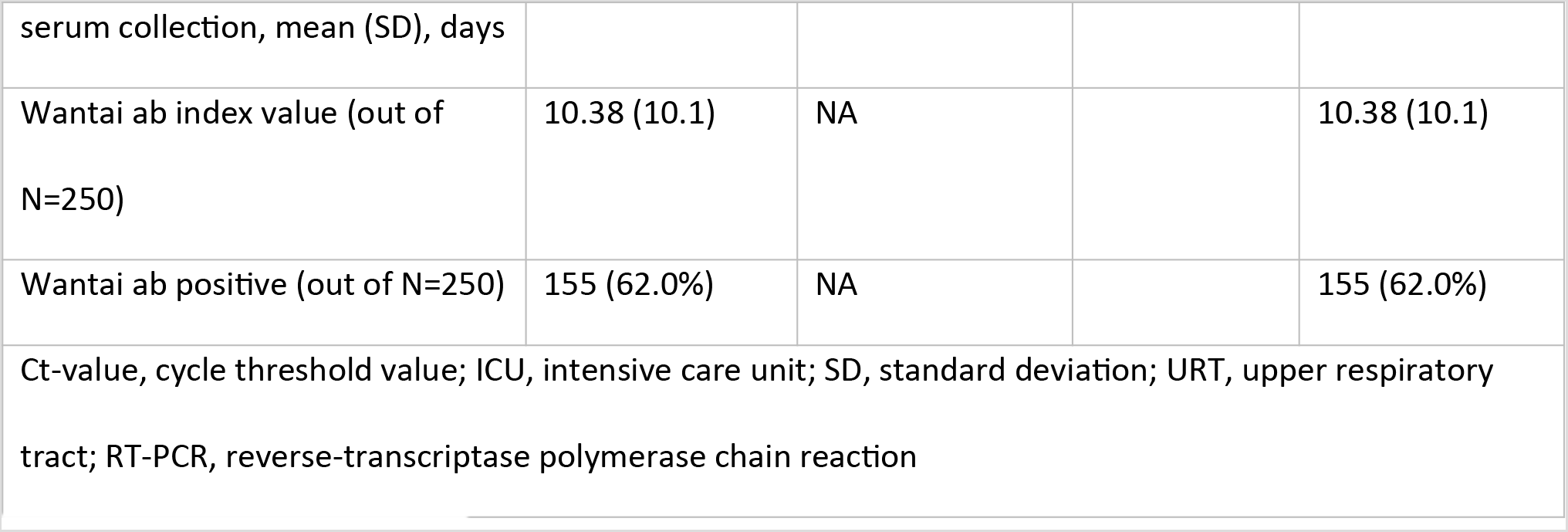
Cohort baseline characteristics

**Figure 1.**
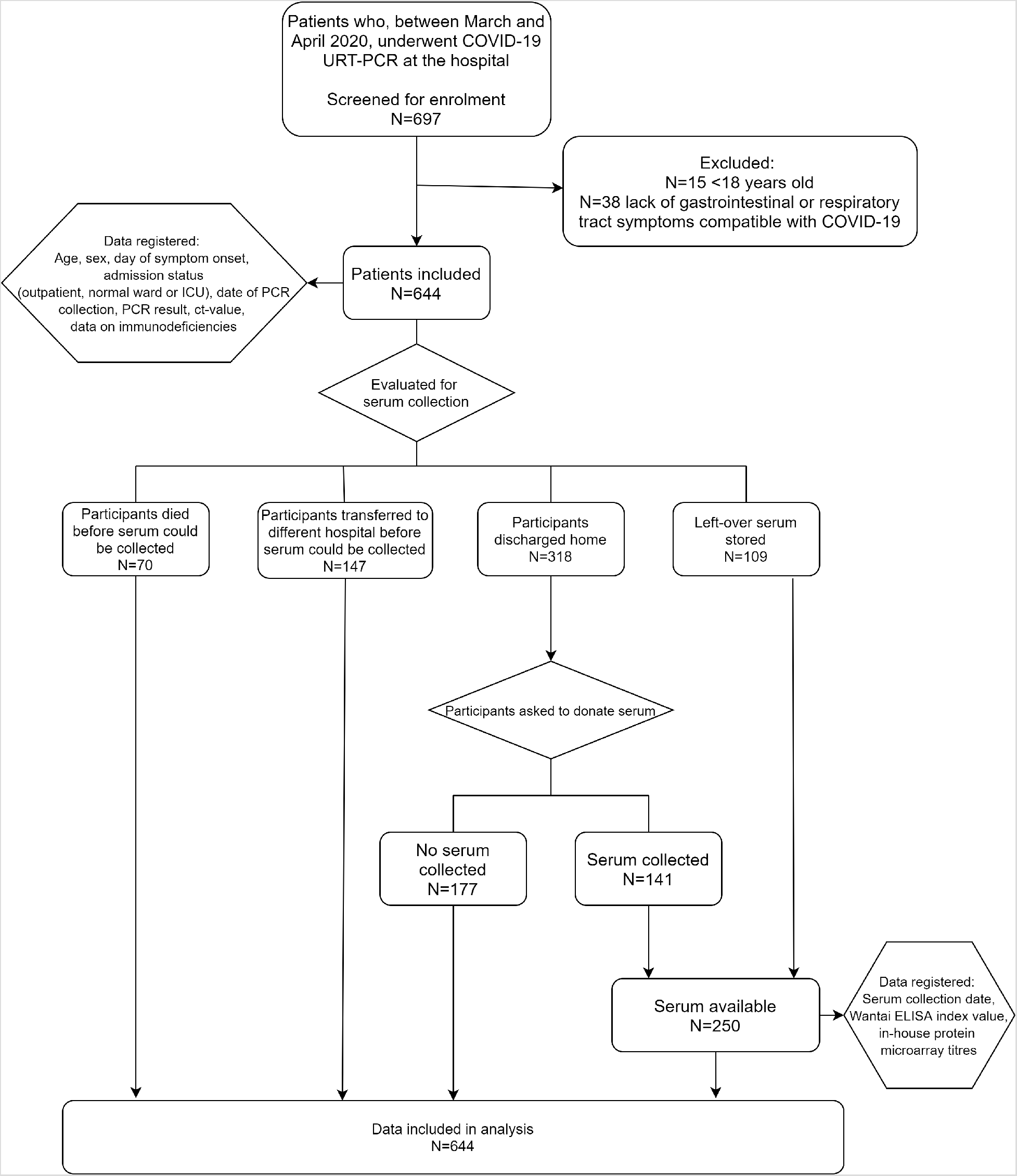
Patient cohort flow-diagram.

**Figure 2.**
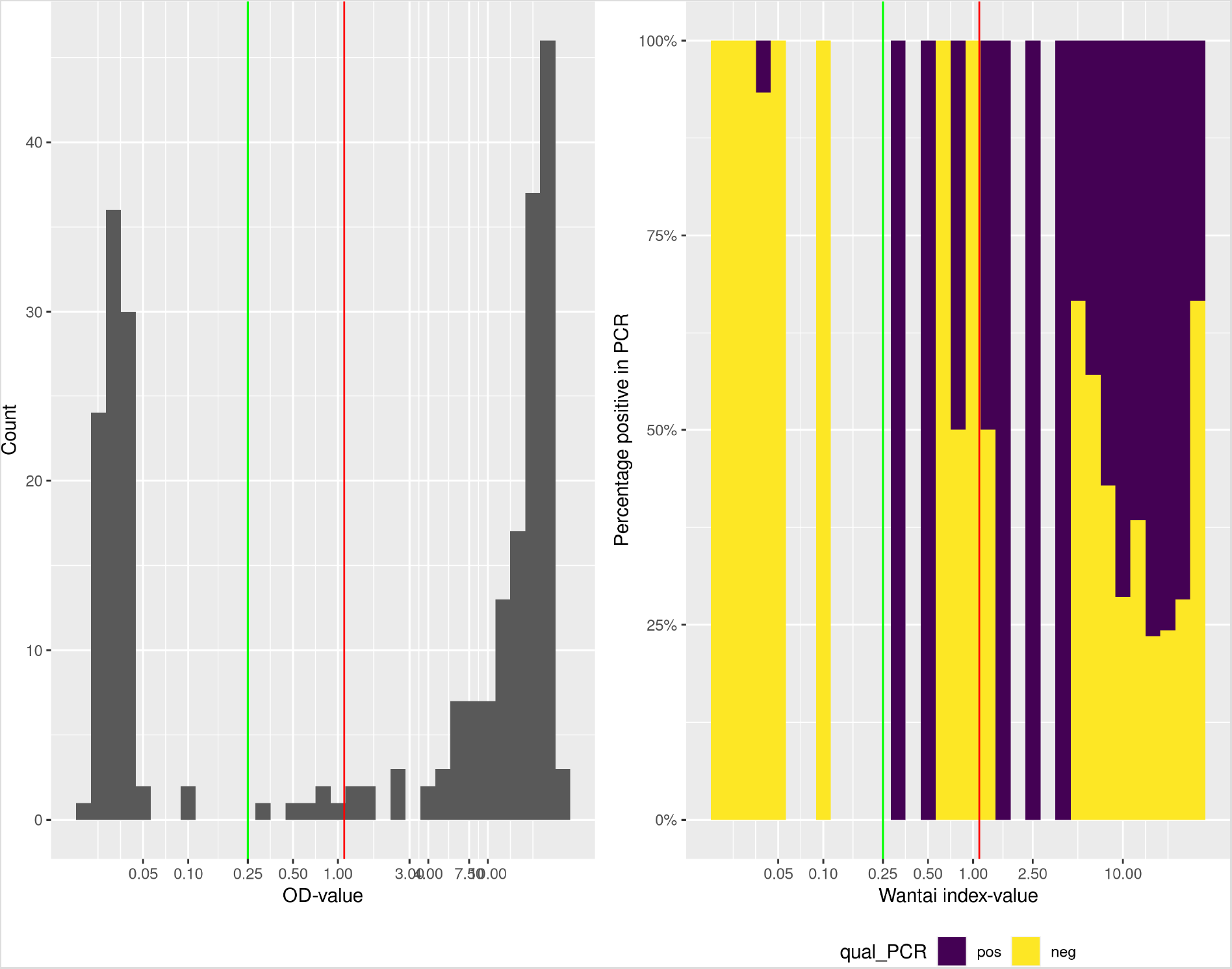
a) Histogram of Wantai total antibody index values on a logarithmic scale. The red vertical line indicates the manufacturer’s cut-off value, the green vertical line our adapted cut-off value. b) Percentage of PCR-positivity within Wantai total antibody index values. The red vertical line indicates the manufacturer’s cut-off value, the green vertical line our adapted cut-off value.

To investigate the sensitivity of RT-PCR on combined OP/NP swabs in clinically suspected COVID-19 patients we applied Bayesian modeling, taking into account all test outcomes at the individual level, without resort to a putative ‘gold standard’. To this end, we exploit that patients received multiple tests, combined with an assumption of perfect specificity, which implies that a single positive test (either ELISA, microarray, or RT-PCR) indicates a positive patient. Perfect clinical specificity can be justified from the high analytical specificity and the fact that patients were clinically diagnosed with COVID-19 symptoms(coughing, sneezing, dyspnea, rhinitis, fever or diarrhea, see Materials and Methods). URTRT-PCR sensitivity varied according to disease severity: for outpatients, 79% (57%-99% Bayesian credible interval (CI)), for patients admitted to a non-ICU hospital ward 86% (77–95 CI), and for patients admitted to ICU 95% (83–1.00 CI) (Fig. 3, Table S1). URT-RTPCR sensitivity was higher in males than in females (91% vs 80%), higher in deceased than in non-deceased patients (95% vs 83%) and higher in immunocompromised than in immunocompetent patients (93% vs 85%) (Fig. S1, Table S1).

**Figure 3.**
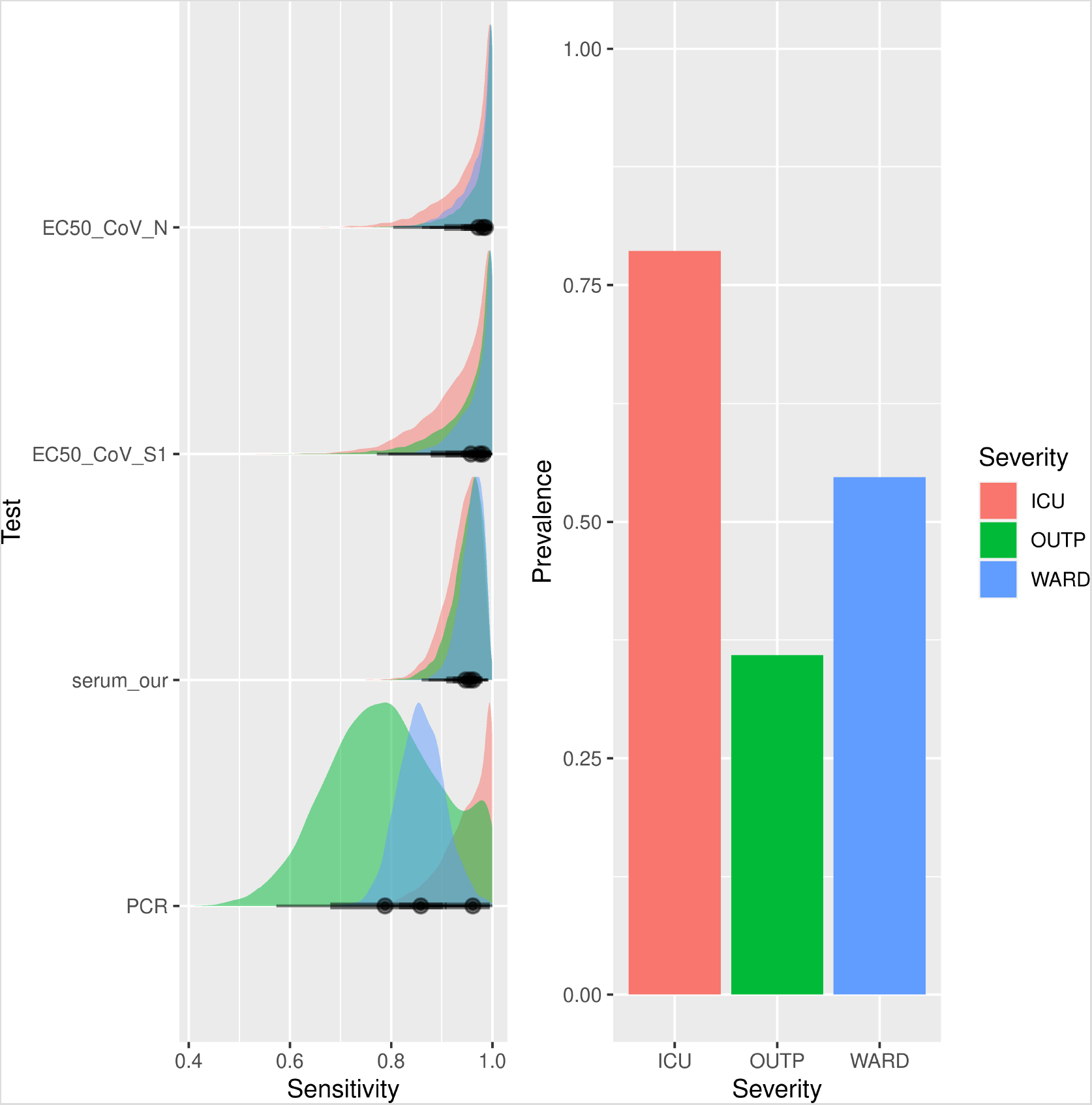
a) Posterior uncertainty distributions of the sensitivities for the microarray assays, serology and RT-PCR assay. Colors indicate severity category. b) Prevalence estimates by severity category. Legend: ICU, Intensive Care Unit; OUTP, outpatients; WARD, non-ICU hospitalized patients; EC50_CoV-N, SARS-CoV-2 N protein microarray; EC50_CoV-S1, SARS-CoV-2 S1 protein microarray; serum_our, Wantai serology using cut-off value of 0.25; PCR, polymerase-chain reaction.

For each disease severity category, URT RT-PCR cycle threshold (Ct)-values displayed an increasing trend by number of days since symptom onset, reflecting a decreasing viral load in time, albeit with considerable inter-individual variation (Fig. 4, Table S2). Ct-values of outpatients were generally higher, even close to symptom onset, and increased rapidly compared to Ct-values of hospitalized patients. Ct-values were not significantly different between males and females, between immunocompromised and immunocompetent patients, or between deceased and non-deceased patients (Fig. S2, Table S2). In patients admitted to a regular ward, URT RT-PCR retained its sensitivity up to at least three weeks post symptom onset (Fig. 5). Clearly, RT-PCR on IC and in-hospital patients retains its initial sensitivity for a prolonged period of time (approximately 15 and 20 days respectively). However, the point of decline is highly uncertain, due to the absence of patient material longer after onset of symptoms. Therefore, conclusions on the time of decline of sensitivity should not be drawn for IC and in-hospital patients. In contrast, outpatients do show a marked decrease of sensitivity over time, with lower uncertainty: sensitivity could be halved already within three weeks. Sensitivities according to sex, mortality and immunosuppression remained stable during at least three weeks post symptom onset (Fig. S3).

**Figure 4.**
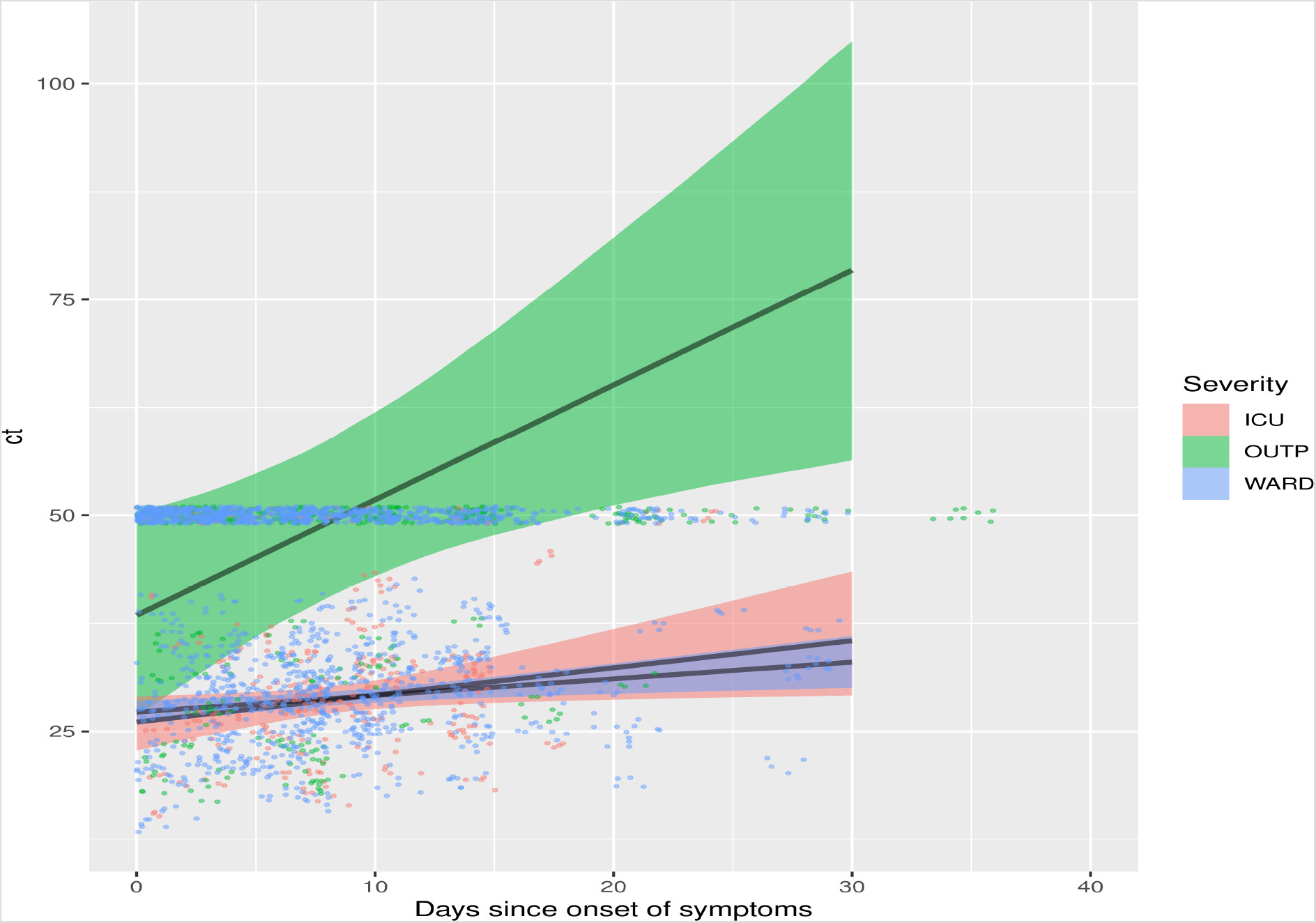
Linear increase of Ct-value in relation to days since onset of symptoms across different disease severity categories. The shaded band indicates 95% Bayesian credible interval. The dots are the original data. Dots positioned at a Ct of 50 were right-censored in the inflated model (i.e. count as either above 50 or a negative individual). Abbreviations: ICU, Intensive Care Unit; OUTP, outpatients; WARD, non-ICU hospitalized patients; ct, Ct-value.

**Figure 5.**
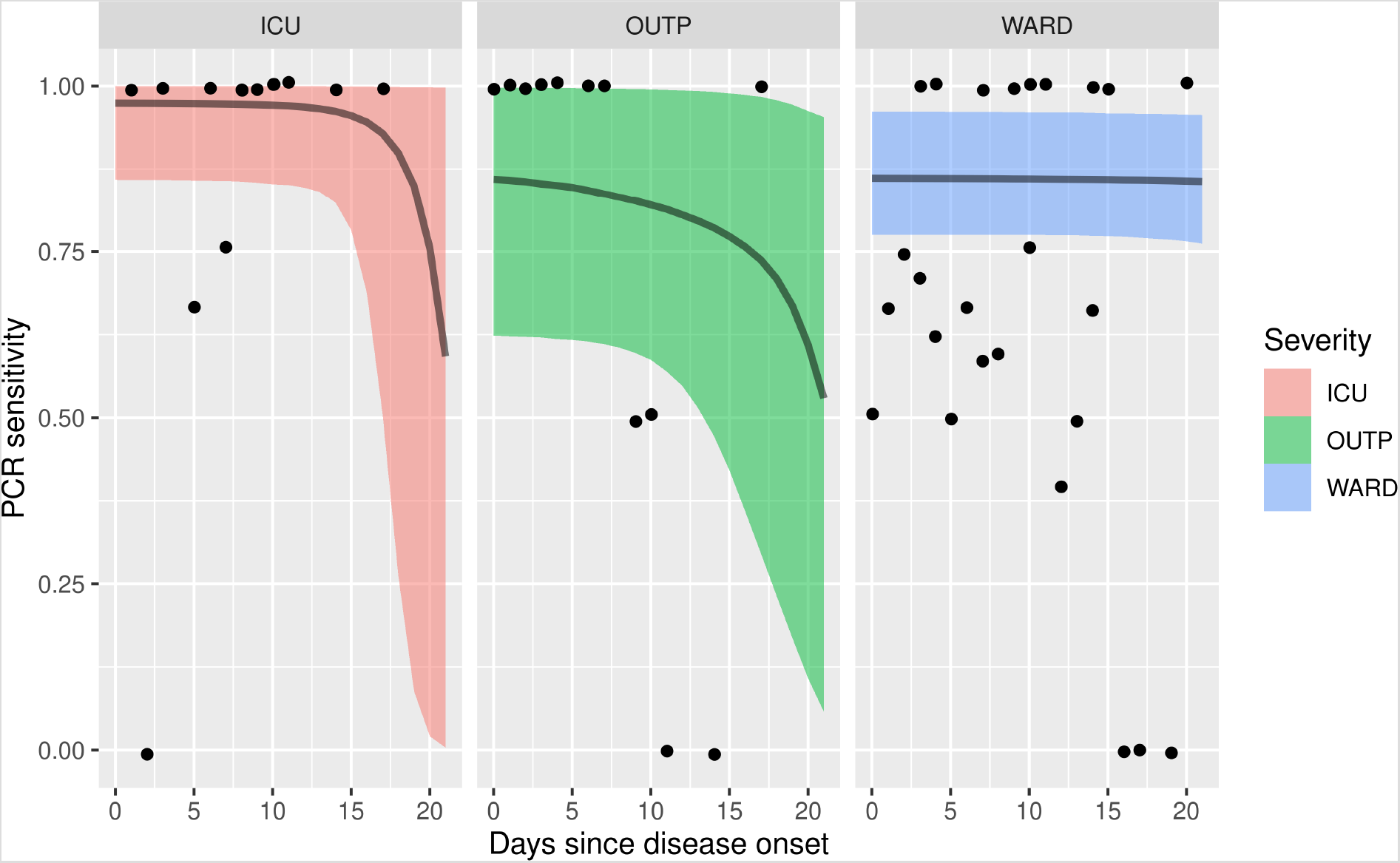
Modeled sensitivities as a function of days since onset of symptoms. Panels and shading indicate disease severity. The bands indicate 95% Bayesian credible interval, and black dots an estimate of sensitivity directly from the original data, by assuming Wantai serology as the gold standard. Abbreviations: ICU, Intensive Care Unit; OUTP, outpatients; WARD, non-ICU hospitalized patients.

COVID-19-suspected patients with a negative URT RT-PCR and without alternative diagnosis were frequently retested for SARS-CoV-2 after resampling. This increased the total number of RT-PCR-confirmed patients with 35 (Table 1). Repeated URT-swab was most commonly used, but had the least added value (10/89, 11.2% positive). Feces generated the most additional positive results (23/58, 39.6%), followed by sputum (6/28, 21.4%). Note that these tests were not included in the Bayesian sensitivity analysis, due to their dependence on the outcome of the initial URT RT-PCR (see Supplementary Text).

Serum samples were analyzed for SARS-CoV-2 antibodies by ELISA and PMA. Bayesian modelling revealed an overall sensitivity of > 94% for both ELISA and PMA, which did not change significantly according to disease severity category, sex or mortality (Fig. 2, Table S1). The sensitivity of antibody assays was lower in immunocompromised patients (82–92%, Table S1) and this was the only group in which antibody assays had a lower sensitivity compared to UTR RT-PCR. The ELISA and PMA results were discrepant in only four of 250 sera (Table S3). All four patients tested RT-PCR positive in URT-swab, and three out of four were immunocompromised.

## Discussion

An accurate assessment of the clinical sensitivity of diagnostic tools, in particular of RT-PCR on URT samples on which global test, trace and isolate strategies are based(*18*), is an absolute requisite for good patient care and adequate infection risk management. We observed a decrease in sensitivity with decreasing disease severity, an increase in sensitivity in immunocompromised patients and a rapid decline of sensitivity in time post onset of symptoms, but only in outpatients. In our study, 5–14% of hospitalized COVID-19 cases and 21% of outpatients tested negative in URT-PCR. This finding is in contrast with local practice and guidelines, which are often based on the assumption of near-perfect sensitivity(*19*).

Frequent false-negatives have been consistently reported in several different countries since the onset of the pandemic. The analytical sensitivity of PCRs generally approaches 100%, which means tests are able to detect a single viral genome copy in the reaction volume(*20*–*23*), the clinical sensitivity may be substantially lower, due to low quality of samples, presence of inhibiting factors, suboptimal pre-analytic processing or specific biologic features of the viral infection. If one assumes infection of the URT always occurs in SARS-CoV-2 infected persons, a negative URT RT-PCR could be the result of localized early clearance of the virus or low levels of local viral replication. The localized clearance would then have different kinetics than the rate of viral load decline we measured in this study. Alternatively, the possibility should be considered that the URT of these patients was not infected and infection can remain more localized (e.g. to the trachea / lower respiratory tract)(*24*). As can be expected, RT-PCR sensitivity of URT correlates with the SARS-CoV-2 RNA load in these samples. The increase of mean Ct-value over time was significantly slower in hospitalized patients compared to outpatients, which explains why the sensitivity of RT-PCR on URT samples decreased more rapidly in cases with mild infections than in hospitalized patients and suggests hospitalized patients have difficulties clearing the virus. In our cohort, immunocompromised patients were the only category in which the sensitivity of serology was lower (PMA), or comparable (Wantai) to URT-RT-PCR. According to the definition we used, 73 of 644 patients were immunocompromised (Table S4). Most patients suffered from conditions severely affecting humoral immunity or were treated by systemic immunosuppressants that inhibit antibody production. This relative lack of antibody response may explain the lower sensitivity of serology in this patient group, and it may clarify why relatively many immunocompromised patients’ Wantai results fell below the manufacturer’s cut-off value. Although antibody assays were shown to have high sensitivity in the rest of our cohort, their use in tracing and isolating strategies is nonexistent, due to the fact that antibodies take a while to be produced and may be detectable for several months after initial infection. Therefore, in order to timely diagnose and quarantine COVID-19 patients, URT-PCR will continue to be used as the assay on which initial decisions are based.

Our estimated URT-PCR sensitivity has important consequences for screening, treatment and isolation measures in hospitals. Though a positive PCR does not necessarily signify the presence of viable virus(*25*), sensitivities of 86–94% are not sufficient to lift isolation measures in the event of a negative initial URT-PCR. In the early stages of hospital admission, clinical suspicion and local COVID-19 prevalence should guide isolation and treatment decisions. Additionally, timely sputum or feces sampling should take place in an effort to confirm COVID-19 diagnosis. Several studies found that 30–50% of COVID-19 patients have detectable SARS-CoV-2 RNA in feces (*26, 27*). In our study, feces yielded 3.5 times more RT-PCR-confirmed positives than repeated URT-swab: 40% of additionally sampled feces tested positive, as opposed to only 11% of repeated URT-swabs (Table 1).

Our findings also have ramifications for broad molecular testing in the general population, as currently established across the world, based on URT-swabbing in high-throughput testing lanes(*28*). The evidently often absent thorough epidemiological and clinical interpretation of negative results in these settings, in combination with the observed low clinical sensitivity of RT-PCR in the population with mild complaints that typically visit those testing sites, will lead to missed cases. For example, if one assumes a sensitivity of 80% of RT-PCR for people with mild symptoms in high-throughput testing lanes and where 200 of 10,000 persons with mild complaints test positive, the apparent prevalence would be 2%. Using the Rogan-Gladen estimator(*29*), the true prevalence would be *Apparent Prevalence/Sensitivity = 2%/80% = 2.5%*. Hence, out of the 9800 persons tested negative, 50 are expected to be false negatives, which the system will fail to isolate.

In conclusion, our results show that for an accurate diagnosis based on RT-PCR test results and subsequent appropriate clinical management and infection control measures, both in hospitals and in public health, a thorough understanding of the clinical sensitivity of RT-PCR in URT samples in clinically heterogeneous patient cohorts is necessary. The apparent lack of a high clinical sensitivity of this standard diagnostic method in specific situations warrants vigilance for missed cases especially in settings of high-throughput testing lanes where epidemiological and clinical context are often disconnected from negative test results for final interpretation.

## Data Availability

Data are available upon request.

## Acknowledgments

We thank BJ Bosch for providing the S1 antigen for the protein microarray and H van Zundert for providing additional data on immunocompromise

## Author contribution

JLM, BJMB, AEN and CBEMR conceived the project; BJMB arranged medical ethical approval, BJMB, AJGvO and AEN provided recruitment of study participants, obtained metadata from COVID-19 patients and obtained clinical samples; YCMK and MB provided additional clinical samples; BJMB and AJGvO coordinated clinical sample processing; GJG and JR processed patient samples; ANS, ABM and BJMB analyzed and interpreted data; BJMB, ANS, JLM, EdV, AEN, CBEMR and JJV wrote and edited the manuscript with input from all listed authors.

## Data and materials availability

The R script and data are available upon request from the corresponding author.

## Ethical approval

The study protocol was approved by both the institutional Science Review Board and the regional Medical Ethics Committee METC Brabant (reference number NW2020–31), with a waiver of informed consent for using left-over serum, drawn for other purposes than this study. Informed consent was obtained from patients who were contacted to have a blood sample drawn for the purpose of this study. The study was performed in accordance with the principles of the Declaration of Helsinki and was not supported by grants of any kind.

